# Airway management, procedural data, and mortality records of patients undergoing surgery for mucormycosis associated with coronavirus disease (COVID-19)

**DOI:** 10.1101/2021.09.06.21263168

**Authors:** Prashant Sirohiya, Saurabh Vig, Tanmay Mathur, Jitendra Kumar Meena, Smriti Panda, Gitartha Goswami, Raghav Gupta, Abhilash konkimalla, Dheeraj Kondamudi, Nishkarsh Gupta, Brajesh Kumar Ratre, Ram Singh, Balbir Kumar, Anuja Pandit, Kapil Sikka, Alok Thakar, Sushma Bhatnagar

## Abstract

**Purpose:** Although unexpected airway difficulties are reported in patients with mucormycosis, the literature on airway management in patients with mucormycosis associated with Coronavirus disease is sparse.

**Methods:** In this retrospective case record review of 57 patients who underwent surgery for mucormycosis associated with coronavirus disease, we aimed to evaluate the demographics, airway management, procedural data, and mortality records.

**Results:** Forty-one (71.9%) patients had a diagnosis of sino-nasal mucormycosis, fourteen (24.6%) patients had a diagnosis of rhino-orbital mucormycosis, and 2 (3.5%) patients had a diagnosis of palatal mucormycosis. A total of 44 (77.2%) patients had co-morbidities. The most common co-morbidities were diabetes mellitus in 42 (73.6%) patients, followed by hypertension in 21 (36.8%) patients, and acute kidney injury in 14 (28.1%) patients. We used the intubation difficulty scale score to assess intubating conditions. Intubation was easy to slightly difficult in 53 (92.9%) patients. In our study, mortality occurred in 7 (12.3%) patients. The median (range) mortality time was 60 (27–74) days. The median (range) time to hospital discharge was 53.5 (10–85) days. The median [interquartile range] age of discharged versus expired patients was 47.5 [41,57.5] versus 64 [47,70] years (P = 0.04), and median (interquartile range) D-dimer levels in discharged versus expired patients was 364 [213, 638] versus 2448 [408,3301] ng/mL (P = 0.03).

**Conclusion:** In patients undergoing surgery for mucormycosis associated with the coronavirus disease, airway management was easy to slightly difficult in most patients. Perioperative complications can be minimized by taking timely and precautionary measures.

## Introduction

Mucormycosis is an acute-onset, aggressive, and rapidly progressive angioinvasive infection caused by saprophytic fungi of the order Mucorales. The most common underlying risk factor associated with mucormycosis is uncontrolled diabetes mellitus. Hematopoietic stem cell and solid organ transplants, corticosteroid therapy, neutropenia, or drug-induced immunosuppression are other identifiable risk factors.[1,2]

There was a sudden increase in cases of mucormycosis in the second wave of Coronavirus disease (COVID-19) in India.[3,4] The management plan of these cases included surgical debridement, systemic antifungal therapy, sugar control, and management of systemic adverse effects related to the antifungal therapy.[5] Diabetes mellitus has been recognized as the most common coexisting concomitant disease. Attention must be given to control blood glucose.[6] In addition, airway management in these patients may be difficult due to the aggressive nature of the disease. The oropharyngeal region may be involved by fungi and edema in the supraglottic region may cause difficult endotracheal intubation and difficult ventilation.[7–9] These patients are receiving injections of amphotericin B which may have significant adverse effects such as nephrotoxicity, hypokalemia, hypomagnesemia, fever, tremor, dyspnea, and hypotension.[10] Apart from this, COVID-19 itself has harmful effects on various organs of the body. Many of these patients require surgical debridement of the involved tissues and anesthesiologists are involved in the multidisciplinary perioperative management of mucormycosis associated with COVID-19.[9]

The mortality rate for mucormycosis associated with COVID-19 is less known but the overall mortality rate for mucormycosis is 54%.[11] There is a risk of involvement of vital structures such as the brain and eye, so surgical debridement should be planned on an urgent basis as a delay can worsen the prognosis. There may be less time to optimize patient comorbidities, making perioperative management challenging. Since all patients in our study are positive for COVID-19, problems caused by wearing personal protective equipment, limited staffing, and supplies are additional difficulties in managing these patients.[12]

Although unexpected airway difficulties are reported in patients with mucormycosis, the literature on airway assessment and management in patients with mucormycosis associated with COVID-19 is sparse. In this study, we aimed to evaluate demographics, airway assessment and management, procedural data, and mortality records in patients undergoing surgery for mucormycosis associated with COVID-19.

## Methods

### Study Design

A retrospective review of the medical records of patients who underwent surgery for mucormycosis associated with COVID-19 at the National Cancer Institute (Jhajjar) between 6 May 2021 and 15 June 2021 was performed.[4] Patients with incomplete medical records were excluded from the study.

### Study Setting and Population

The study included 57 COVID-19 positive patients who underwent surgery for mucormycosis under general anesthesia. The study was approved by the Institute Ethics Committee (IEC-450/02.07.2021) of the All India Institute of Medical Sciences, New Delhi.

### Study Objectives

The primary objective was to describe airway assessment and management and the secondary objective of the study was to describe demographics, procedural data, and mortality records.

### Study Protocol

Preoperative data included demographic characteristics, disease type, patient comorbidities, smoking and alcohol status, laboratory investigations, vital parameters, and airway assessment. The airway was assessed under the following headings: mouth opening, presence of loose teeth, dentures or bucktooth, whether the patient was edentulous, Mallampatti scoring, neck movements, thyromental distance, presence of short neck, presence of receding chin, and post-surgical changes.[13,14] All the surgeries were performed under general anesthesia. All anesthesiologists involved in the procedure had at least 3 years of clinical experience. As a protocol, if mouth opening was adequate, we used a conventional C-MAC video laryngoscope for endotracheal intubation. We calculated the Intubation difficulty score (IDS) to assess intubating conditions.[15,16] This includes the following parameters: number of intubation attempts, number of operators, number of alternative techniques, Cormack-Lehane grade, whether lifting pressure is necessary, whether or not laryngeal pressure is applied, and vocal cord mobility. IDS value = 0 represents easy intubation, a score of >0 and ≤5 represents slightly difficult intubation, and a score >5 represents moderate to major difficult intubation. The time required for intubation and the percentage of glottic opening (POGO) were also noted.[17] Data related to the procedure were also recorded in terms of nasogastric tube insertion, invasive monitoring, central venous catheter insertion, blood loss, urine output, etc.

The duration of anesthesia was defined as the time between induction of anesthesia and transfer of the patient to the post-anesthesia care unit (PACU) or ICU. Patients who required postoperative mechanical ventilation or who had undergone extensive surgery were transferred directly from the operation theater to the ICU. Patient data regarding length of hospital stay and mortality were obtained from hospital records. Duration of hospital stay was defined as the number of days between the date of hospitalization to the date of discharge/death.

### Statistical Analysis

Data related to the variables selected in the study were extracted from the records and entered into MS Excel software version 16.0 (Microsoft Inc.) for summative analysis. Data were summarized using the median with the interquartile range [25th, 75th] or median (range) for continuous variables and numbers and proportions (%) for categorical variables. Statistical review of the data was performed using IBM SPSS version 24 (SPSS Inc., Chicago IL, USA). A P-value of less than 0.05 was considered statistically significant.

## Results

The demographic and clinical characteristics of the patients are presented in Table 1. The 57 patients consisted mostly of men 35 (61.4%) with a mean age of 49 (26,78) years. The median body weight and height were 64 (46,82) kg and 1.67 (1.45–1.81) meters. Most patients had American Society of Anesthesiologists (ASA) scores II (40.4%) and III (40.4%).[18] Forty-one (71.9 %) patients had sino-nasal mucormycosis, fourteen (24.6%) had rhino-orbital mucormycosis (out of which 2 patients had a cerebral extension), and 2 patients (3.5%) was diagnosed with palatal mucormycosis. Total 44 (77.2 %) patients had co-morbidities. The most common co-morbidity was Diabetes Mellitus 42 (73.6%), followed by hypertension 21 (36.8%) and Acute kidney injury 14 (28.1%).

**Table 1:** Demographic and clinical characteristics of included cases in the study.

|  |  |
| --- | --- |
| **Variable** | **Frequency (%) / Median [IQR]** |
| 1. Age (Years) | 49 [41.5,59.5] |
| 2. Weight (Kilograms) | 64 [58,70] |
| 3. Height (meters) | 1.67 [1.58,1.71] |
| 4. Sex |  |
| Male | 35 (61.4) |
| Female | 22 (38.6) |
| 5. Diagnosis |  |
| Sino-nasal | 41 (71.9) |
| Rhino-orbital | 14 (24.6) |
| Palatal | 2 (3.5) |
| 6. Clinical Outcome |  |
| Discharge | 50 (87.7) |
| Expired | 7 (12.3) |
| 7. ASA Grade |  |
| 1 | 11 (19.3) |
| 2 | 23 (40.4) |
| 3 | 23 (40.4) |
| 8. Comorbidity |  |
| Diabetes | 42 (73.7) |
| Hypertension | 21 (36.8) |
| Acute Kidney Injury | 14 (28.1) |
| Chronic Kidney Disease | 2 (3.8) |
| Hypothyroidism | 2 (3.8) |
| Coronary artery disease | 2 (3.8) |
| Asthma | 1 (1.8) |
| 9. Smoking Status* | 10 (17.5) |
| 10. Alcohol intake Status* | 10 (17.5) |
| 11. Pre-operative Pulse Rate (per minute) | 89 [79.5,100.5] |
| 12. Pre-operative Systolic Blood Pressure | 125[117.5,136.5] |
| 13. Pre-operative Diastolic Blood Pressure | 75 [66.5,84.5] |
| 14. Pre-operative Respiratory rate (breaths per minute) | 22 [20,24] |

*\* Items are mutually non-exclusive*

Airway assessment and management, and procedural data parameters are presented in Table 2. In all 57 patients, mask ventilation was without difficulty. Fifty-six (98.3%) patients underwent oral endotracheal intubation using a conventional C-MAC laryngoscope. One patient had restricted mouth opening (<1 finger), for which we performed nasotracheal intubation using a fiberoptic bronchoscope. The first pass intubation success rate was 92.9 %.

**Table 2.** Airway Assessment and management with Procedural Data.

| **Table 2 Airway Assessment and management with Procedural Data** |  |
| --- | --- |
| **Variable** | **Frequency (%)** |
| 1. Mouth opening |  |
| >3 Fingers | 49 (85.9) |
| 2.5-3 Fingers | 3 (5.3) |
| 2-2.5 Fingers | 4 (7.0) |
| <1 Fingers | 1 (1.8) |
| 2. Loose tooth | 5 (8.8) |
| 3. Artificial tooth | 3 (5.3) |
| 4. Edentulous | 2 (3.5) |
| 5. Buck tooth | 2 (3.5) |
| 6. Mallampatti Score |  |
| 1 | 3 (5.3) |
| 2 | 44 (77.2) |
| 3 | 8 (14.0) |
| 4 | 2 (3.5) |
| 7. Neck Movements | 56 (98.2) |
| 8. TMD (adequate) | 46 (80.7) |
| 9. Short neck | 7 (12.3) |
| 10. Receding chin | 2 (3.5) |
| 11. Post-surgical changes | 0 (0) |
| 12. No. of attempts |  |
| 1 | 53 (92.9) |
| 2 | 4 (7.0) |
| 13. No. of operators |  |
| 1 | 56 (98.2) |
| 2 | 1 (1.8) |
| 14. Cormack Lehane grade |  |
| 1 | 24 (42.9) |
| 2 | 28 (50.0) |
| 3 | 4 (7.0) |
| 15. Lifting force required |  |
| Normal | 24 (42.9) |
| Increased | 32 (57.1) |
| 16. Vocal cord mobility |  |
| Abduction | 57 (100) |
| 17. External laryngeal maneuver | 21 (36.8) |
| 18. Esophageal intubation | 0 (0) |
| 19. Intubation Stylet used | 2 (3.6) |
| 20. Intubation difficulty score (n=56) |  |
| 0 | 11 (19.6) |
| 1 | 13 (23.2) |
| 2 | 17 (30.4) |
| 3 | 11 (19.6) |
| 4 | 1 (1.8) |
| 5 | 3 (5.4) |
| 21. Extubated | 41 (71.9) |
| 22. Shifted to ICU (Intubated) | 16 (28.1) |

We calculated the Intubation difficulty score to assess intubating conditions and we found a score of 0 (Easy) in 11 patients, a score of >0 to ≤5 (slightly difficulty) in 42 patients, and a score >5 (moderate to major difficulty) in 3 patients.

The median percentage of glottic opening (POGO) score was 80 (20,100)% and the time required for intubation was 15 (10,180) seconds. All patients received crystalloids while 2 patients additionally received blood transfusions. We performed peripherally inserted central catheterization in 47 (82.5%) patients and internal jugular vein cannulation in 7 (12.3%) patients. In 3 patients, internal jugular venous cannulation was performed before their surgery. Nasogastric tube insertion was performed in 19 (33.3%) and Foley catheterization was performed in 19 (33.3%) patients. The median blood loss was 150 (50,500) ml and urine output was 250 (200,300) ml. Preoperative investigations are listed in Table 3. The median duration of anesthesia was 150 (90,270) min. Forty-one (71.9%) patients were extubated after surgery in the operation theatre. Sixteen (28.1%) patients were transferred to the intensive care unit for postoperative elective mechanical ventilation. The reason for postoperative elective mechanical ventilation was extensive surgery in 14 patients and inadequate reversal from anesthesia in 2 patients. Eight out of 16 patients were extubated on postoperative day 0, 6 patients on postoperative day 1, and 2 patients on day 2. After extubation, a high flow nasal cannula (HFNC) is required in one patient for 10 days. The patient was later weaned from it.

**Table 3.** Pre-operative Investigations of patients.

| **Table 3 Pre-operative Investigations of patients** |  |
| --- | --- |
| **Variable** | **Median [IQR]** |
| 1. Pre-Op Blood sugar (mg/dL) | 146 [122.5,171] |
| 2. D-dimer (ng/mL) | 407 [227,657.75] |
| 3. Fibrinogen (mg/dL) | 484 [415.75,556] |
| 4. Prothrombin Time (sec) | 11.8 [10.97,12.97] |
| 5. INR | 1 [0.93,1.14] |
| 6. Hemoglobin (g/dL) | 11.35 [10.32,12.7] |
| 7. WBC Count (/μL) | 8.84 [7,11.94] |
| 8. Platelet Count (/μL) | 235.5 [181.25,303.75] |
| 9. Neutrophils (%) | 77.3 [69.5,85.05] |
| 10. Ferritin (ng/mL) | 862.85 [495.62,1381.57] |
| 11. Lactate dehydrogenase (units/L) | 295 [225,349] |
| 12. C-reactive protein (mg/L) | 11.22 [5.02,14.99] |
| 13. Procalcitonin (ng/mL) | 0.09 [0.04,1.17] |
| 14. Interleukin-6 (pg/mL) | 17.5 [8.7,43] |
| 15. Total Bilirubin(mg/dL) | 0.43 [0.3,0.58] |
| 16. Direct Bilirubin(mg/dL) | 0.18 [0.1,0.26] |
| 17. Alanine aminotransferase (U/L) | 29 [17,45] |
| 18. Aspartate Aminotransferase (U/L) | 24 [20.5,35] |
| 19. Total Protein(g/dL) | 5.9 [5.31,6.4] |
| 20. Albumin (g/dL) | 3.2 [2.8,3.6] |
| 21. Albumin/Globulin Ratio | 1.27 [1.07,1.43] |
| 22. Alkaline Phosphatase (IU/L) | 98 [87,138.5] |
| 23. Urea (mg/dL) | 34.2 [21.2,48.15] |
| 24. Creatinine (mg/dL) | 0.87 [0.69,1.7] |
| 25. Calcium (mg/dL) | 8.11 [7.87,8.64] |
| 26. Sodium (mmol/L) | 136 [134,139] |
| 27. Potassium (mmol/L) | 4 [3.55,4.7] |
| 28. Uric Acid (mg/dL) | 4.3 [3.3,5.1] |

In our study, mortality occurred in 7 (12.3%) patients. The median (range) mortality time was 60 (27–74) days. The median (range) time to hospital discharge was 53.5 (10–85) days. We did a comparative analysis between discharged patients and expired patients. The median [interquartile range] age of discharged versus expired patients was 47.5 [41,57.5] versus 64 [47,70] years (P = 0.04), and median (interquartile range) D-dimer levels in discharged versus expired patients was 364 [213, 638] versus 2448 [408,3301] ng/mL (P = 0.03).

## Discussion

Surgical debridement of mucormycosis is an invasive procedure and airway management of patients can be challenging. Involvement of the oropharyngeal region by fungus and supraglottic edema can lead to difficulties with mask ventilation and endotracheal intubation.[8] In one study, 3 patients had fungal debris in the oropharyngeal region, and in one of these patients, due to supraglottic edema, a video laryngoscope was used for endotracheal intubation.[19] In our study, mask ventilation was not difficult in any of our patients. First pass intubation success was 92.9%. Two attempts were made in 4 (7.0%) patients. Two patients required an intubation stylet for their endotracheal intubation and two patients had difficulty guiding the endotracheal tube through the glottis on the first attempt but the second attempt was successful. In one patient, restricted mouth opening was present, therefore, fiberoptic bronchoscope-guided intubation was performed. Rest all patients are intubated by C-MAC video laryngoscope as per departmental COVID-19 protocol. We did not find any fungal debris in the oropharyngeal region and supraglottic edema in any of our patients. The intubation difficulty score was calculated with the help of the intubation difficulty scale. Eleven patients had no intubation difficulty, 42 patients had slight intubation difficulty (score 1–4), 3 patients had moderate to major intubation difficulty (score >5).

We had a dedicated COVID-19 operation theatre with all the healthcare staff wearing level 3 personal protective equipment.[20] We checked all equipment and drugs for anticipated or unanticipated difficult airway management. We had masks of different sizes, stylets, bougies, laryngeal mask airways of different sizes, video laryngoscope (C-MAC) blades of different sizes, fiberoptic bronchoscope, 2 working suction apparatus, and an emergency tracheostomy trolley for management of the unexpected difficult airway.

COVID-19 is a major contributor to morbidity and mortality due to dyspnea, poor functional status, chest pain or tightness, hypercoagulability, endocrine abnormalities especially impaired glycemic control, etc. This, in addition to mucormycosis, makes the prognosis worse in these patients.[21] In our study 42 (73.7 %) patients had a history of diabetes which was managed by administration of insulin perioperatively. Central venous catheters may be required peri-operatively for blood or blood product transfusions, for fluid replacement, inotropic or vasopressor support, and long-term infusion of amphotericin B.[22] We used Groshong® PICC catheter (4 and 5 Fr) in 47 patients and internal jugular venous cannulation in 7 patients. The use of systemic amphotericin B in the management of mucormycosis associated with COVID-19 has its distinct toxicities, the most important being nephrotoxicity. There are other side effects of amphotericin B such as hypokalaemia, hypomagnesemia, fever, dyspnoea, shivering, and hypotension.[23] In our study, 14 patients had pre-operative amphotericin B-induced nephrotoxicity.

The primary objective of surgical management is to debride all necrotic tissues. Our patients received both antifungal treatment and surgical treatment. The overall mortality rate of mucormycosis is 54%.[11] The mortality rate of mucormycosis associated with COVID-19 is still unknown. In our study, mortality occurred in 7 (12.3%) patients. The median mortality time was 60 (range, 27-74) days. The median (range) mortality time was 60 (27–74) days. The median (range) time to hospital discharge was 53.5 (10–85) days. Five (12.2%) of 41 patients with sinonasal mucormycosis and 2 (14.3%) of 14 patients with rhino orbital mucormycosis (cerebral extension in 1 patient) expired. None of the patients with palatal mucormycosis expired.

Our study has some limitations. First, the study had a retrospective nature and was based on analysis of anesthesia and hospital records that may be subject to selection bias. Second, all the patients were from a single center. The number of expired patients was comparatively small (7 patients) for statistical comparison with those who were discharged (50 patients).

## Conclusion

In patients undergoing surgery for mucormycosis associated with the coronavirus disease, airway management was easy to slightly difficult in most patients. Perioperative complications related to airways, the effect of COVID-19 and diabetes mellitus, systemic effects of amphotericin B can be reduced by taking timely and precautionary measures.

## Data Availability

Data will be available on request

